# Integration of Electrochemical Sensing and Machine Learning to Detect Tuberculosis via Methyl Nicotinate in Patient Breath

**DOI:** 10.1101/2024.05.23.24307746

**Authors:** Mary A. Jeppson, Zachary Rasmussen, Robert Castro, Talemwa Nalugwa, Esther Kisakye, Wilson Mangeni, Alfred Andama, Devan Jaganath, Adithya Cattamanchi, Swomitra K. Mohanty

**Affiliations:** Department of Chemical Engineering, The University of Utah, Salt Lake City, UT 84112 USA; Division of Pulmonary and Critical Care Medicine, University of California San Francisco, San Francisco, CA 94110 USA; Walimu, Kampala Uganda; Walimu and the Department of Medicine, Makerere University College of Health Sciences, Kampala, Uganda; Division of Pediatric Infectious Diseases and the Center for Tuberculosis, University of California San Francisco, San Francisco, CA 94110 USA; Division of Pulmonary Diseases & Critical Care Medicine, University of California Irvine, Irvine, CA 92697 USA and the Center for Tuberculosis, University of California San Francisco, San Francisco, CA 94110 USA; Department of Chemical Engineering and the Department of Materials Science and Engineering, The University of Utah, Salt Lake City, UT 84112 USA

**Keywords:** electrochemical sensor, machine learning, point-of-care detection, triage test, square wave voltammetry, volatile organic biomarker, XGBoost

## Abstract

Tuberculosis (TB) remains a significant global health issue; making early, accurate, and inexpensive point-of-care detection critical for effective treatment. This paper presents a clinical demonstration of an electrochemical sensor that detects methyl-nicotinate (MN), a volatile organic biomarker associated with active pulmonary tuberculosis. The sensor was initially tested on a patient cohort comprised of 57 adults in Kampala, Uganda, of whom 42 were microbiologically confirmed TB-positive and 15 TB-negative. The sensor employed a copper(II) liquid metal salt solution with a square wave voltammetry method tailored for MN detection using commercially available screen-printed electrodes. An exploratory machine learning analysis was performed using XGBOOST. Utilizing this approach, the sensor was 78% accurate with 71% sensitivity and 100% specificity. These initial results suggest the sensing methodology is effective in identifying TB from complex breath samples, providing a promising tool for non-invasive and rapid TB detection in clinical settings.

## Introduction

The World Health Organization’s Global Tuberculosis Report revealed that approximately 1.3 million people died of tuberculosis (TB) in 2023, and is the leading cause of death by a single infectious agent [1]. Although often treatable, many people lack access to diagnosis, resulting in an estimated one-third of TB cases going unreported [1]. In high-income countries, the time from symptom onset to treatment initiation averages 25 days, while in low-income countries, the time to treatment averages 56 days. Numerous healthcare provider visits and diagnostic tests are often required before patients receive a diagnosis [1]. Additionally, drug-resistant TB has emerged as a growing health concern since 2020, increasing morbidity, mortality, and the urgency for prompt treatment to prevent disease transmission [1].

To address this public health emergency, the World Health Organization (WHO) launched the “End TB” initiative, aiming to reduce the global average time from TB symptom onset to treatment to less than one month by 2025. A key factor in achieving this goal is improving diagnostic access. However, current diagnostic methods, including molecular testing, require extensive laboratory infrastructure and are expensive and time-consuming to administer [1].

Researchers are now focusing on developing affordable, non-sputum, biomarker-based, point-of-care devices, aiming to meet the WHO target product profile for a diagnostic at ≥66% sensitivity and ≥98% specificity, or a triage test reaching ≥90% sensitivity and ≥70% specificity [2]. There is growing interest in developing diagnostics that detect volatile organic biomarkers (VOBs) in breath, as collection is non-invasive and pulmonary TB is the most common type of TB disease. While current commercial products show promise, they have yet to meet the WHO’s sensitivity and specificity requirements and are not targeted for the detection of specific VOBs [1].

Traditionally, techniques such as gas chromatography-mass spectrometry (GC-MS) have been used to identify and correlate VOBs to disease states [3]. Portable sensors can be designed by modeling electrochemical reactions between a biomarker and a metal salt solution containing a supporting electrolyte and a transition metal, using approaches like dynamic functional modeling (DFT) [4],[5]. Ray et al. demonstrated that four TB biomarkers, methyl phenylacetate, methyl p-anisate, methyl nicotinate and o-phenyl anisole; selectively bind with transition metals, particularly cobalt (II) and copper (II), allowing for the specific detection of electrochemical signals associated with these biomarkers [5]. However, further refinement of detection methods is needed in order to meet the WHO’s sensitivity and specificity targets for TB diagnostics.

FWe previously reported the successful detection of the TB biomarker methyl-nicotinate (MN) using copper as the electroactive solution (EAS) [6]. MN is semi-volatile and thus liquid phase detection, rather than gas phase detection, is preferable [7]. This approach offers more consistent results due to the presence of a counter electrode, which reduces the impact of production variability from the working electrode [7]. For VOB detection, square wave voltammetry (SWV) is an electroanalytical technique that applies a series of square wave potentials to an electrode, resulting in improved sensitivity and selectivity [8]. This method detects trace amounts of analytes, such as VOBs, while minimizing background interference, making it a promising approach for TB diagnosis [8]. The result is a unique electrochemical pattern for a given analyte. Machine learning can then be utilized to automate detection of these patterns for disease classification.

In this work, we applied a square wave voltammetry method using Copper (II) metal salts to detect MN from adults being evaluated for pulmonary TB in Uganda. We then applied a machine learning model, specifically XGBoost, to correlate the extracted features from the electrochemical data with the TB status of the patients.

## Methods

An experimental flow of the research is shown in Figure 1.

**Fig. 1.**
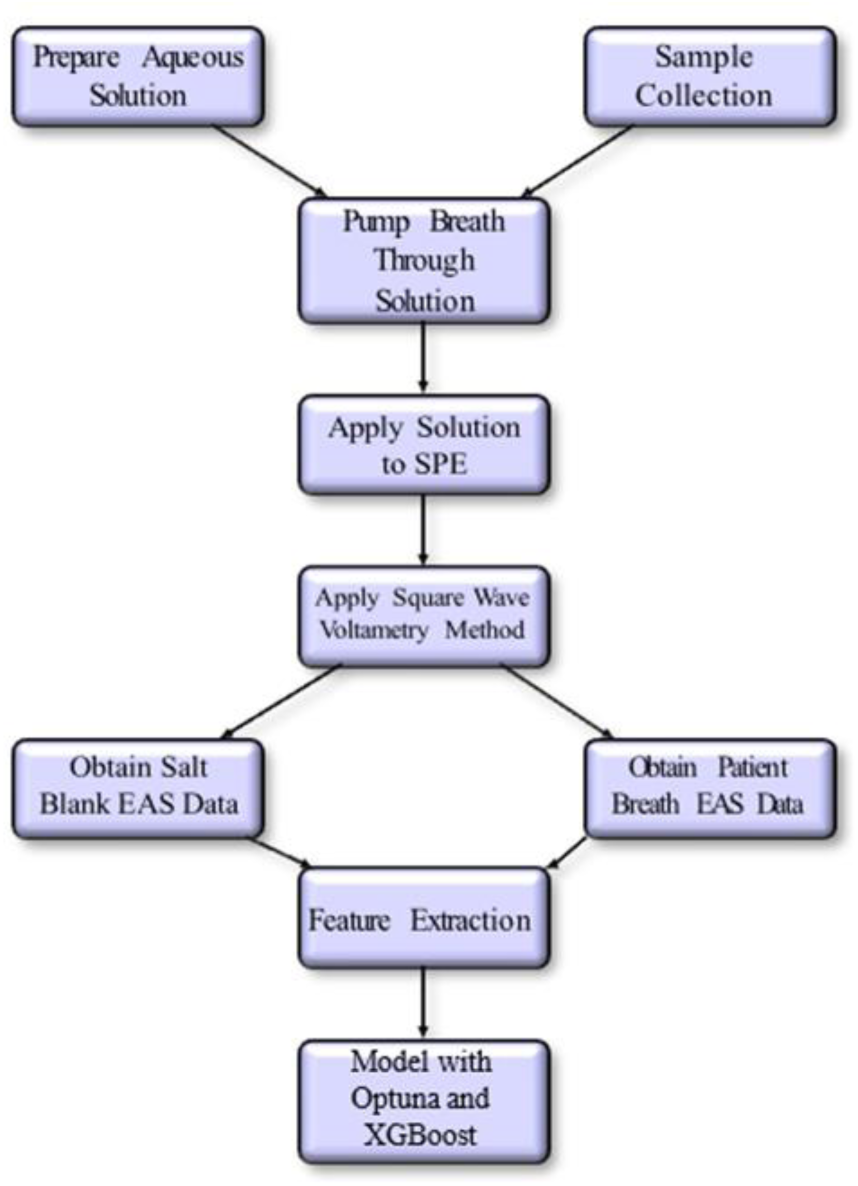
Flow chart for data acquisition, feature extraction, and modeling in XGBoost with Optuna to optimize hyperparameters.

### A. Preparation of Aqueous Solutions

Aqueous solutions were prepared using deionized water (≥ 18MΩcm−1 type 1 ultrapure, PURELAB Classic). Base electroactive solutions consisted of aqueous copper(II) chloride (Alfa Aesar, anhydrous, 98% min) as the active metal and sodium chloride (Fisher Chemical, ≥ 99.0%) as the supporting electrolyte with concentrations of 1mM copper chloride and 100mM sodium chloride.

### B. Preparation of Spiked Breath Mimics Containing MN

A stock solution containing 10 mM of methyl-nicotinate (MN) (Sigma-Aldrich ≥ 99.9%) dissolved in deionized water was prepared for examining the effects of known concentrations of MN in mimic breath samples. Spiked mimics were created by collecting 24 healthy breath samples in 10 liter Tedlar bags. The breath was then transferred to the metal salt solution using the breath transfer device using a flow rate of 1L/min. The MN stock solution was diluted and added to the solution to achieve a concentration of 0.1mM MN in the resulting spiked breath solutions.

### C. Collection and Processing of TB Patient Breath Samples

We analyzed breath samples from 57 adults from Kampala, Uganda, who presented to care at Mulago National Referral hospital and Kisenyi Health Centre IV with at least 2 weeks of cough and were evaluated for TB with sputum-based molecular testing (Xpert MTB/RIF Ultra, Cepheid, Sunnyvale) and mycobacterial culture. Of the 57 participants, 42 (74%) had microbiologically confirmed TB, and 15 (26%) had negative microbiological TB testing. Participants were asked to exhale into 10-liter Tedlar® Breath Analysis Bags (CEL Scientific), which were then collected and pumped through the copper salt solution at a rate of approximately 1 L/min. This breath-solution transfer device was designed to eliminate sample contamination by pulling gas from the bag into the solution. This was accomplished by placing the pump at the end of the flow path after the breath sample had been pulled into the salt solution, as seen in Figures 2 and 3.

**Fig. 2.**
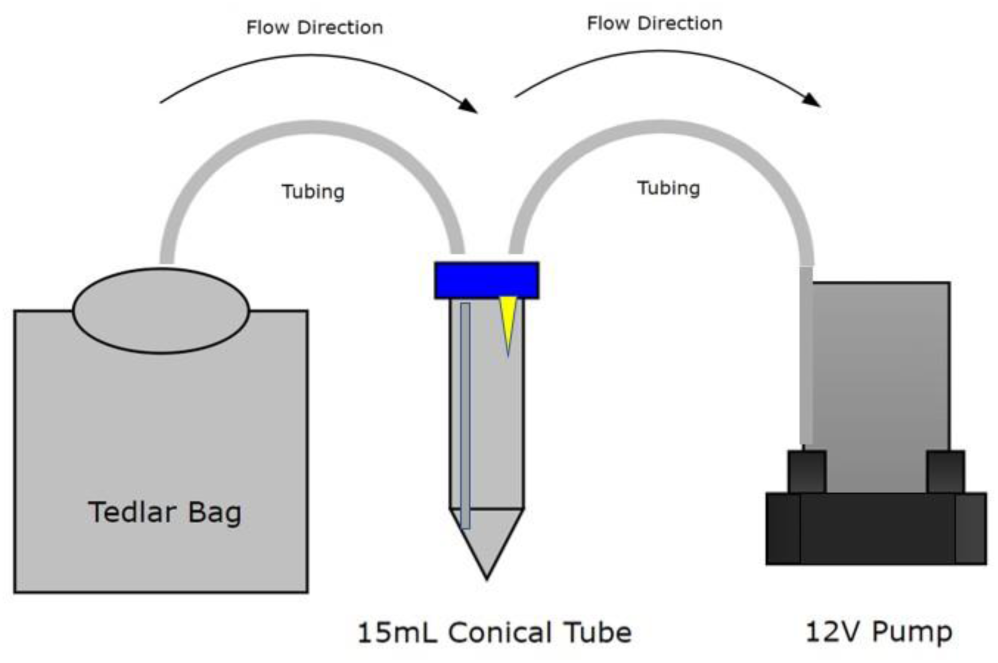
Breath transfer device schematic showing breath flow path. The device was specifically designed to avoid sample contamination from the pump interior by pulling the sample directly into the transfer device.

**Fig. 3.**
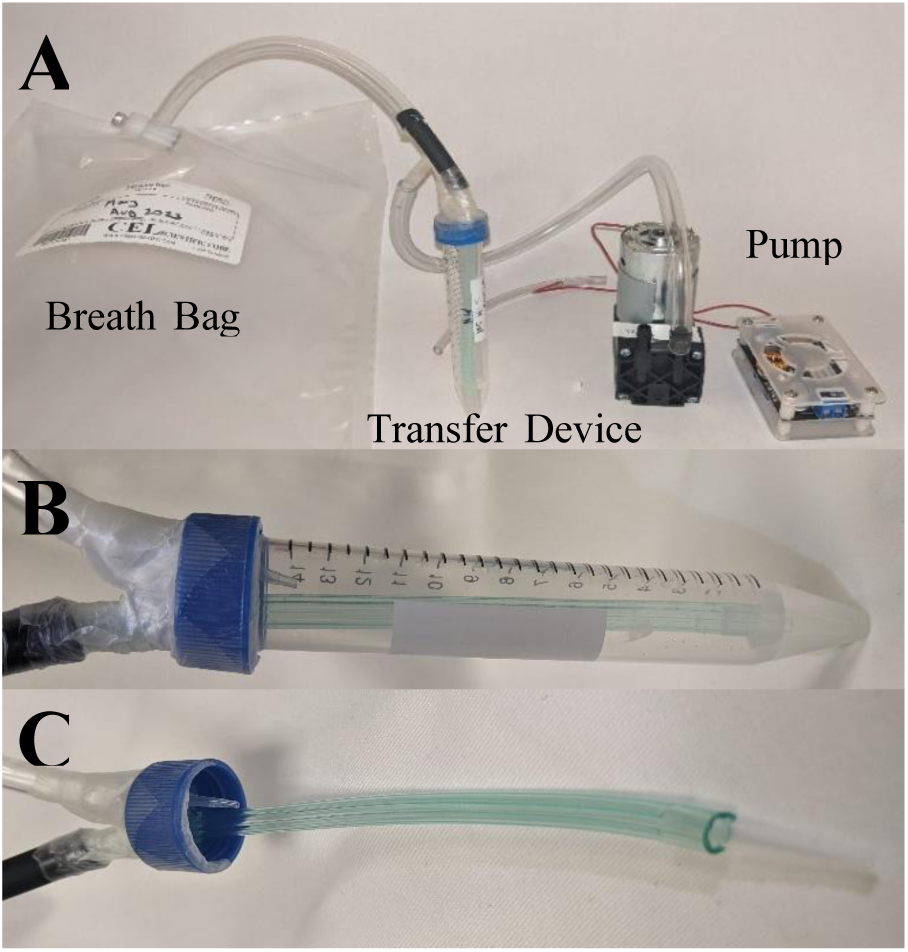
Images of the Breath Transfer System: (A) Full System, (B) Transfer Device, (C) Transfer Device Tubing

### D. Electrochemical Analysis of Spiked Mimics and Clinical Breath Samples

Liquid solutions were electrochemically analyzed using a hand-held potentiostat (PalmSens, EmStat) and screen-printed electrodes (SPE) with unmodified carbon working/counter electrodes and a silver reference electrode (DRP110, 4mm diameter WE, Metrohm-DropSens). The salt solution with dissolved breath for the spiked mimics and clinical samples was applied to the SPE surface with a volume of 150µL. A SWV method was then run with the settings seen in Table I.

**TABLE I.** Method Settings.

| Parameter | Value |
| --- | --- |
| t equilibration | 5 seconds |
| E begin | -0.5 V |
| E end | 0.4 V |
| E step | 0.005 V |
| Amplitude | 0.025 V |
| Frequency | 2 Hz |

### E. Data Analysis and Feature Extraction

Key peaks for redox reactions between MN and Cu were previously identified using SWV from our prior work [6]. Utilizing spiked samples containing 0.1mM MN, we examined the changes in more detail and measured differences between daily blanks and clinical samples. Features associated with redox reactions between MN and copper in the spiked mimics were identified and normalized to the salt blank by assigning a percentage to each current value relative to the max current in the daily salt blank.

The peak features observed in the SWV were normalized and extracted, and the data was split into training and test sets, with 75% of the data used for training and 25% used for validation. We applied an Extreme Gradient Boosting (*i.e.,* XGBoost) model to the training data, a gradient-boosted machine learning algoriothm ideal for handling imbalanced data sets like ours. The XGBoost model parameters were optimized using Optuna, an automatic hyperparameter optimization software as seen in Table II [7]. Python 3.6.9, with the XGBoost, Scikit learn and Optuna packages were used.

**TABLE II.** Best hyperparameters for accuracy.

| Hyperparameter | Value |
| --- | --- |
| Max Depth | 3 |
| Gamma | 0.1368553692983 |
| Min Child Weight | 1 |
| Learning Rate | 0.0149452437099 |
| Subsample | 1.0 |
| n estimators | 81 |
| Colsample Bytree | 0.5 |
| Reg Alpha | 1.1973540541e-06 |
| Reg Lambda | 6.4888499897e-06 |
| Seed | 3 |

These values were found by conducting 50 trials and minimizing the loss function for each. In the independent test set, we evaluated the performance of the trained XGBoost model by calculating the Area under the ROC (Receiver Operating Characteristic) curve (AUC-ROC), and then sensitivity and specificity metrics with 95% confidence intervals (CIs).

## Results

An examination of the current response of the spiked breath mimics, showed a distinct ‘Shoulder peak’ at a low concentration of MN (0.1 mM) along with three other peaks (Figure 4). These features represent specific oxidation/reduction reactions that occur for Cu under SWV as published in our prior work [6]. For example, Peak 1 is theorized to represent Cu^2+^ ↔ Cu^+^, Peak 2 is Cu^2+^ ↔ Cu^0^ and Peak 3, Cu^+^ ↔ Cu^0^. We observed in our prior work and in our breath mimics that the current associated with these peaks vary in relation to MN concentration, with reductions in relative current values in both the ‘Shoulder Peak’ and ‘Peak 2’ exceeding 25% and 20%, respectively [6]. While the mimic data was used to guide the feature extraction process, this data was not used to train or test the patient data.

**Fig. 4.**
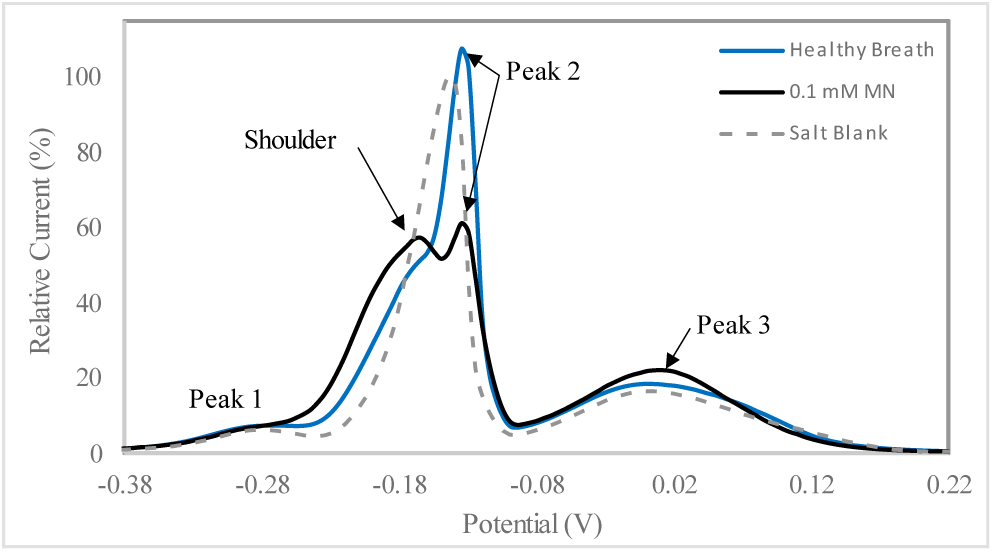
Healthy breath and spiked breath with 0.1 mM MN added. No shoulder present in healthy breath and current reduction is <20%. When MN is added to the breath, a shoulder appears and the relative current values of shoulder and peak 2 are >25% and >20%, respectively.

Similar current behavior was seen in the breath samples of Ugandan participants. The majority of samples were from people with Confirmed TB (74%). A consistent reduction in current was seen for Peak 2 as well as a reduced ‘Shoulder’ current value in participants with confirmed TB relative to symptomatic participants without TB. Figures 5-7 show typical responses for Ugandan participants with and without TB normalized to the maximum current in a daily salt blank. TB negative patients tended to display either the response seen in Figure 6, where no ‘Shoulder’ could be seen, or a response with a ‘Shoulder’ but little relative current reduction as seen in Figure 7. The relative current values for peaks 1,2, and 3 as well as the Shoulder value when present were then extracted from the training data set as features for the XGBOOST model.

**Fig. 5.**
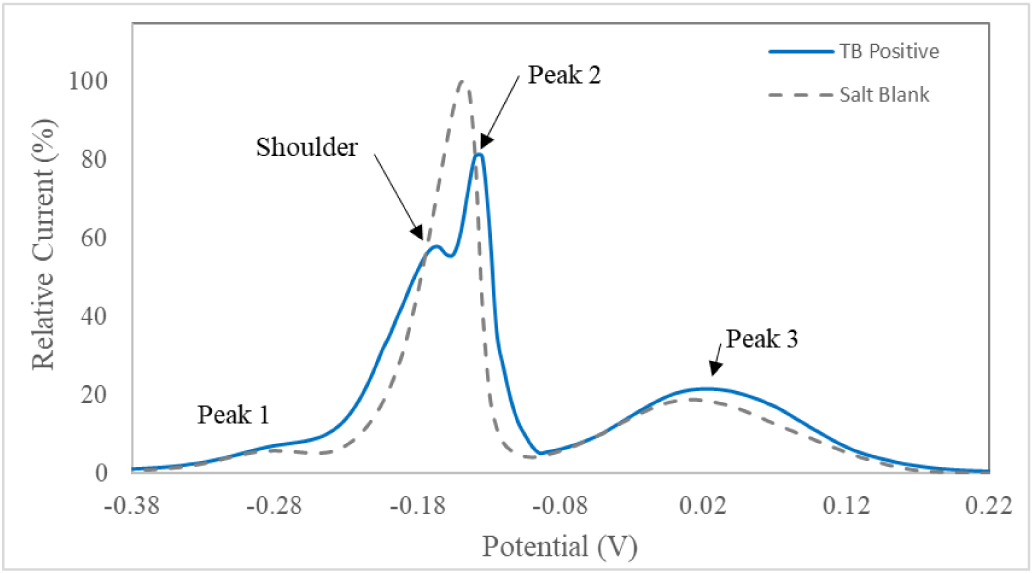
TB-Positive patient example. Shoulder present. Current reduction is <25% and *<*20% at the shoulder and peak 2 apexes, respectively, relative to the maximum current of the salt blank.

**Fig. 6.**
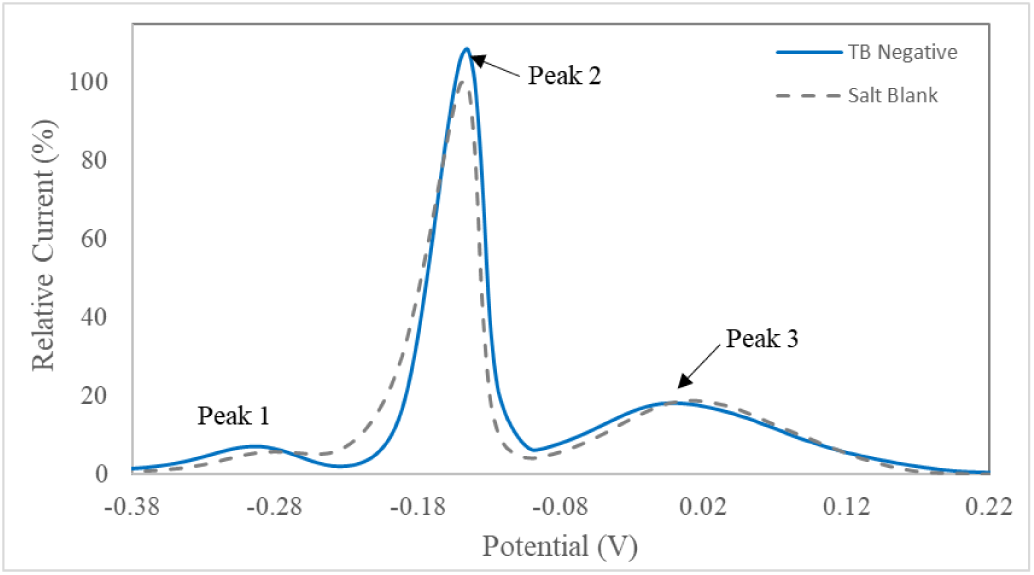
TB-negative patient example. No shoulder peak present. Peak 2 apex current values reduced by *<* 20% relative to the maximum current of the salt blank.

**Fig. 7.**
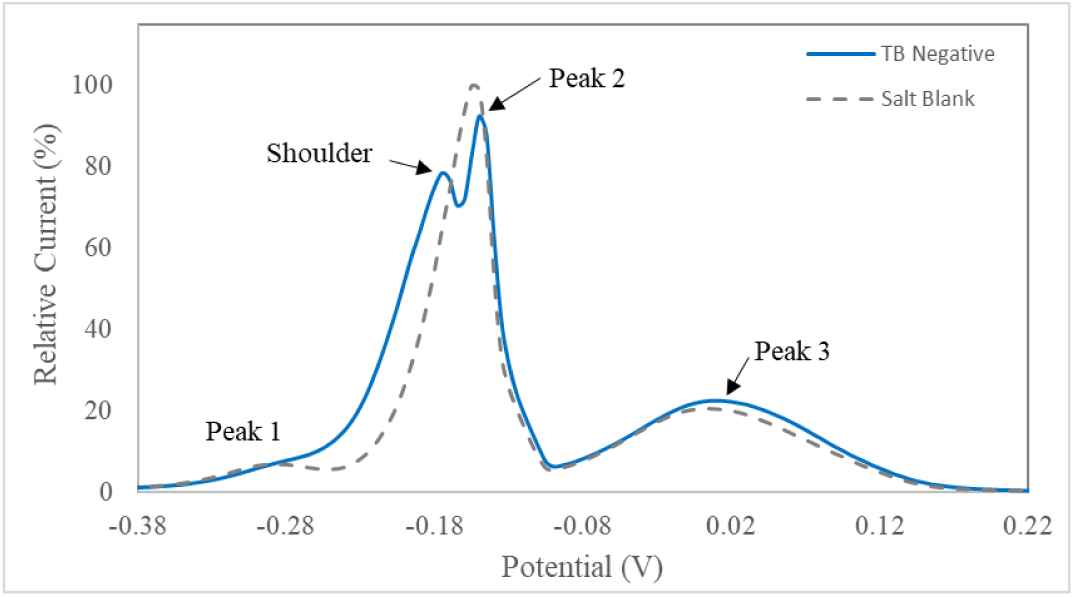
TB-negative patient example. Shoulder present but the current reduction is <25% and <20% at the shoulder and peak 2 apexes, respectively, relative to the maximum current of the salt blank

In comparison to a microbiological reference standard, the sensor response was shown to have an accuracy of 77.8% in the test set, and an area under the curve (AUC) value of 0.964 as shown in the receiver operator characteristic (ROC) curve in Figure 8. In addition to the AUC, we also computed sensitivity and selectivity for both the training and test data, seen in Table III.

**Fig. 8.**
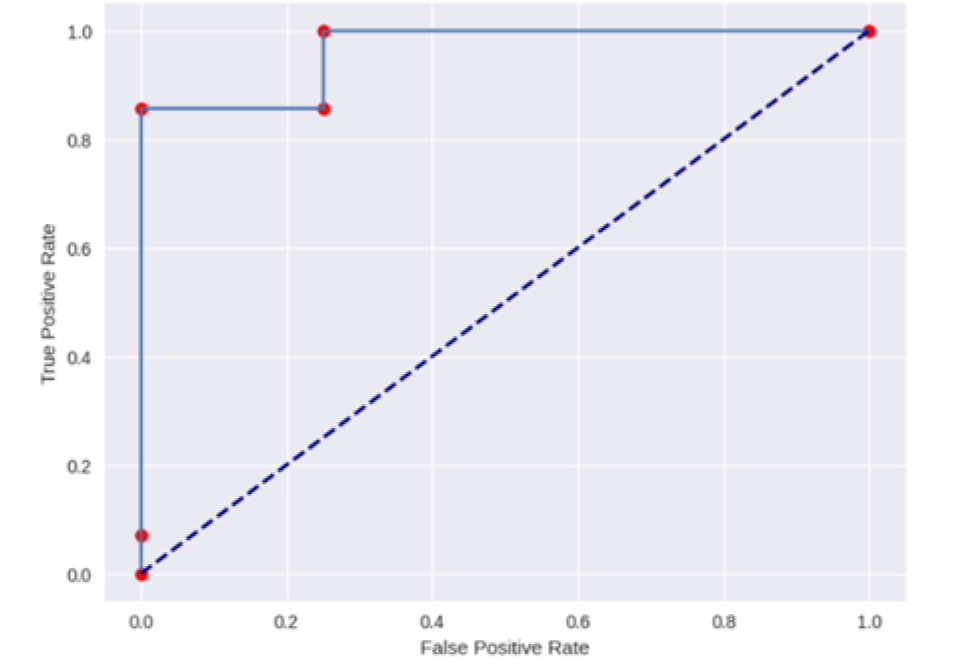
XGBoost Testing data ROC results. Sensitivity and specificity were found to be 0.7143 and 1.000, respectively.

**TABLE III.**
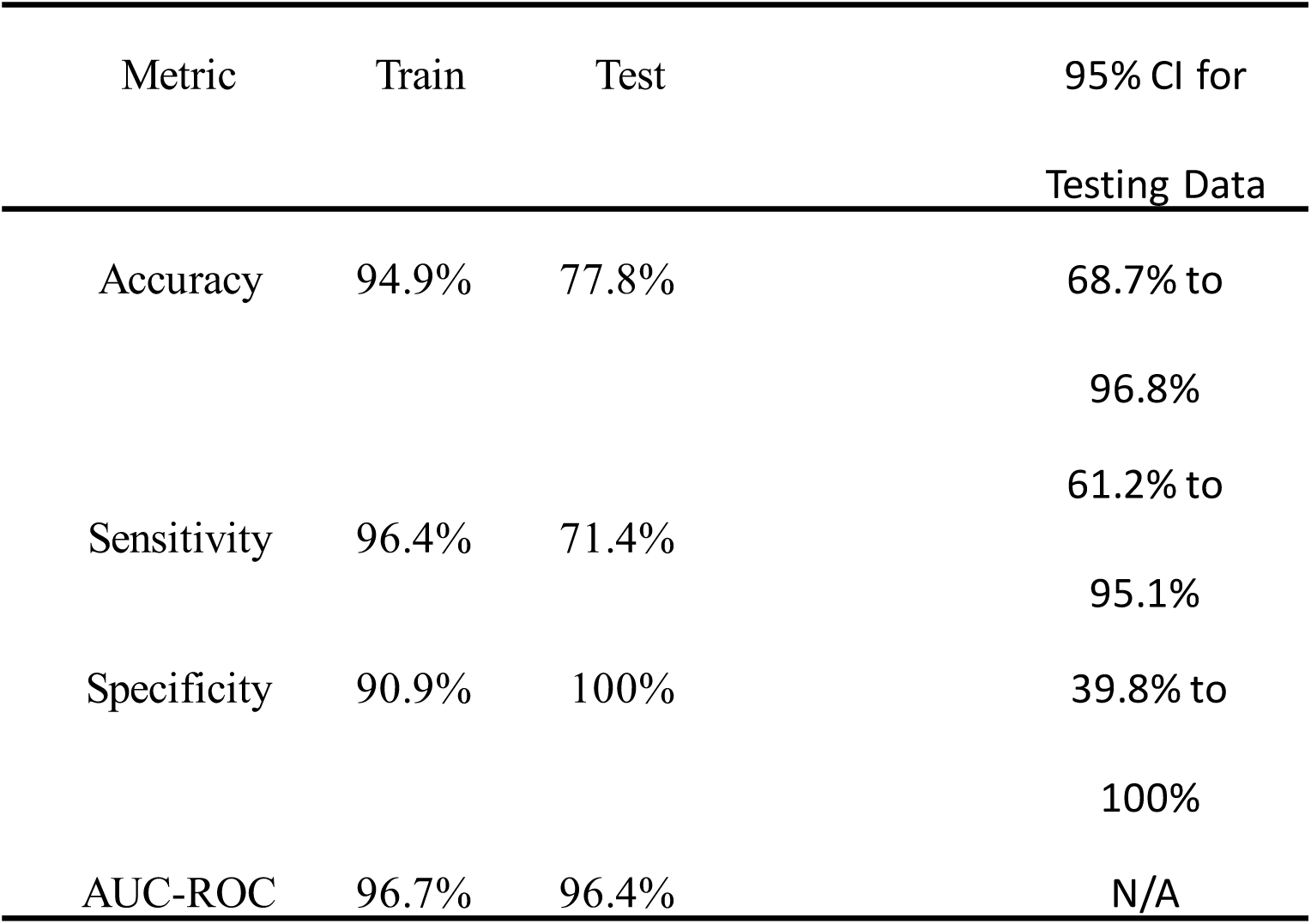
Performance metrics for the optimized model.

For the training data, the model achieved a sensitivity of 96.4% and a specificity of 90.9%. When applied to the testing data, the model yielded a sensitivity of 71.43% (95% CI 61.2% to 95.1%) and a specificity of 100% (95% CI 39.8% to 100%). The XGBoost feature importance analysis results showed that Peak 1 appeared to have the greatest impact on status classification followed by Peak 2 as seen in Figure 9.

**Fig. 9.**
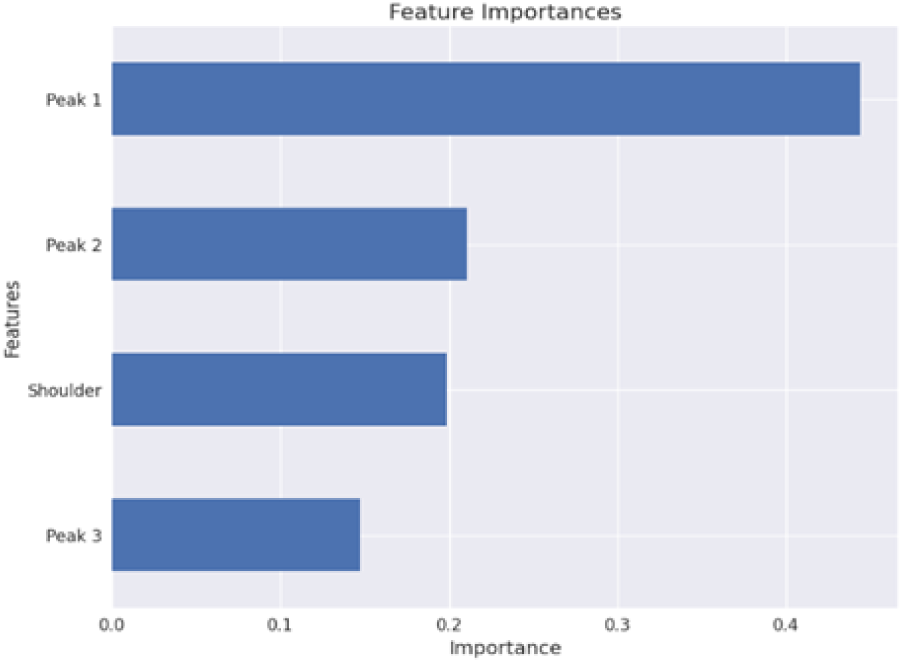
Feature Importance defined by XGBoost algorithm. The Peak 1 feature was assigned the greatest weight in sample categorization, followed by Peak 2.

## Discussion

This study demonstrates the potential of square wave voltammetry (SWV) with our copper(II) EAS sensing platform for the detection of MN, a TB biomarker, in breath samples. Given these initial results, this clinical demonstration shows the applicability of this approach to advancements in TB disease diagnostics, with promising performance metrics that approach the WHO’s target product profile for TB diagnostic tests. These results suggest that this electrochemical technique may have the potential to detect MN at the range of concentration levels typically present in the breath of TB patients.

Human breath contains large amounts of VOCs in complex mixtures and biomarkers related to TB are expected to appear in relatively low amounts, thus limiting the applicability of GC-MS analysis for diagnostic purposes [8]. Our method circumvents this difficulty by leveraging the ability of MN to preferentially dissolve in aqueous solution and tailoring our metal salt solution and electrochemical method to produce a detectable elec trochemical response even at low concentrations.

Our breath mimics spiked with low concentrations of MN validate the response seen in our patient data by showing similar current responses.

Xpert MTB/RIF, the initial test for TB as recommended by the WHO for all people exhibiting TB symptoms [9], requires expensive equipment [10] and sputum samples. However, sputum can be difficult or impossible for some populations to produce, including children and people living with the human immunodeficiency virus (PLWH) [11]. Therefore, the WHO recommended the development of non-sputum based testing. In this context, tests that use samples such as breath or urine are being prioritized. A comparison of approaches that utilize these samples is shown in Table IV.

**TABLE IV:** Summary of Emerging TB diagnostic approaches.

| TEST | SENS.<br>(95% CI) | SPEC.<br>(95% CI) | POC<br>OR<br>LAB | SPECIFIC<br>BIOMARKER | SAMPL<br>E<br>TYPE | TIME-TO-RESULT |
| --- | --- | --- | --- | --- | --- | --- |
| EAS MN | 71.4%<br>(61.2% to<br>95.1%) | 100%<br>(39.8% to<br>100%) | POC | yes | breath | <10<br>minutes |
| ALERE<br>LAM | 18-42%[14]<br>(31%-<br>55%[17]) | 95%<br>(89% to<br>96%) | POC | yes | urine | 25 min[14] |
| E-NOSE | 52.3-76.5%[13]<br>(39.6% to<br>64.7%) | 36.4%[13]<br>(12.4% to<br>68.4%) | POC | no | breath | <1 min[18] |
| GC-MS | 71.2%[12] | 72%[12] | Lab | Yes | breath | >60<br>minutes |

EAS-MN: Electroactive Solutions Methyl Nicotinate ALERE-LAM: ALERE- Lipoarabinomannan E-NOSE: Electronic Nose GC-MS: Gas Chromatography Mass Spectroscopy

The technology presented in this work can potentially provide an early, rapid, and cost-effective diagnosis to millions who otherwise would not receive treatment. Relative to other non-sputum TB testing methods, this approach costs a few dollars, uses well established electrochemical methods, is portable using low-cost instrumentation (< $50), and requires no specialized equipment and training to operate. Current results with the limited number of patients show a sensitivity of 71.4% (CI: 61.2-91.5%) and specificity of 100% (CI: 39.8-100%). To compare, GC-MS analysis of breath with Monte-Carlo simulations has been used to correlate biomarkers of TB [12] and in a study of 251 patients, 130 with active pulmonary TB, the approach achieved a sensitivity of 71.2% and a specificity of 72.0%. However, the cost of one GC-MS system is over $100,000 and requires infrastructure making it not useful in point-of-care applications.

In other breath based approaches using e-nose (Diagnose, C-it BV), Bruins et al. conducted a validation study of 148 participants. While sensitivity and specificity was high for classifying breath profiles between healthy and TB samples, 95.9% and 98.5%, respectively; these values fell to 76.5% and 97.2% when distinguishing participants with TB relative to the full testing population [15]. Recent search on the Diagnose, C-it BV platform shows no development as of 2024. Another study in 2019, used an advanced eNose device to detect pulmonary TB in 287 patients after calibrating the device with 182 individuals. They found sensitivity and specificity values of 78% and 42%, respectively, in the validation phase [16]. When compared to the EAS approach, E-nose platforms are expensive and use a non-targeted VOC detection approach to produce a full-spectrum electronic signal that is fed into an algorithmic classifier [13]. While interesting, currently the lack of using a targeted biomarker contributes to their lower performance.

Finally when comparing the EAS sensing approach to urine based tests such as AlereLAM, the test falls short in sensitivity at 18% in people without HIV and 42% in PLWH according to two studies [14].

While the initial results of our study are promising, limitations include our small sample size and TB classification imbalance. The small sample size also contributes to model overfitting, as seen with the drop in performance from training to test set, although we sought to mitigate this risk by reducing the ratio of features used (Colsample Bytree), the number of estimators, and the max depth. As such, our sensitivity and specificity metrics should be taken as preliminary results. As the cohort expands, more data and class balance will improve model training and validation estimates.

In order to enhance the sensitivity of the method, further investigation of possible biomarker amplification in the SWV method is needed. As seen in our feature examples above, the incompletely resolved feature identified as the ‘Shoulder’ in our evaluation metrics appeared to have a significant impact on the performance of the model predictions. We theorize that a separate reaction with MN may be occurring here. Further resolution of the ‘Shoulder’ from both ‘Peak 1’ and ‘Peak 2’ could provide a fuller characterization of this possible reaction. Possible approaches to this could be leveraging the various solubilities of breath compounds to eliminate those that produce competing reactions in the salt solution. By modifying these processes, the electrochemical signal can be optimized to detect lower concentrations of TB biomarkers, potentially improving diagnostic accuracy. As noted in Figure 9, Peak 1 appears to be most significant. Peak 1 is theorized to represent Cu^2+^ ↔ Cu^+^ which represents a very unstable valent state of Cu, making it less likely to occur and may explain why the peak is typically lower in comparison to the others. Peak 1 may indicate specific interactions between MN and copper which may be diagnostically significant.

Future work will involve systematic studies exploring signal amplification, a detailed investigation of the Shoulder Peak phenomenon, and specific reactions that contribute to Peak 1. Understanding the electrochemical reactions responsible for this peak could provide valuable insights into the sensor’s reaction mechanism; guiding the development of more selective and sensitive diagnostic tools. Further studies will also include the collection of more samples to validate the model, as well as the exploration of other machine learning models and feature engineering techniques to enhance performance.

## Conclusions

This study demonstrates that our SWV method combined with a Copper (II) EAS sensing platform shows promise as a point-of-care diagnostic test for detecting pulmonary tuberculosis biomarkers in patient breath. In future work, these preliminary results will be built upon by further optimization; particularly in method parameters; and exploring ways to boost biomarker concentration in the EAS solution. More work is needed to understand the electrochemical processes driving peak formation and widening the sensor’s range to detect more TB biomarkers. With further refinement of our approach, we will move closer to achieving the global priority of creating a rapid, non-invasive, and cost-effective point-of-care device for TB diagnosis.

## Data Availability

All data produced in the present study are available upon reasonable request to the authors

## Acknowledgements

This work was supported by the National Institutes of Health, National Heart, Lung and Blood Institute, under Grant R01HL139717, and the National Institutes of Allergy and Infectious Diseases, under Grant U01AI152087.

## Notes

### Competing Interest Statement

The authors have declared no competing interest.

### Author Declarations

Makerere University, College of Health Science School of Medicine, IRB Approval Granted University of California-San Francisco Human Research Protection Program Institutional Review Board IRB Approval Granted IRB # 20-32670

### Summary of Updates

This version of the manuscript as been updated to include acknowledgement of funding sources.

